# Capturing additional variance in impact of adolescent depression from free text: proof of concept and preliminary results

**DOI:** 10.64898/2026.11.02.26367115

**Authors:** Anthony Y. Matar, Juan Antonio Lossio-Ventura, Francisco Pereira, Dylan M. Nielson

**Affiliations:** Machine Learning Core, National Institute of Mental Health, NIH

## Abstract

Depression in adolescents has grown in prevalence in the past decade. Despite indications of differences in symptom presentation, measurement properties, and treatment response between adolescents and adults, research is dominated by adult participants. Qualitative work has identified symptomatic and functional domains important to depressed adolescents and those around them; however, there remains little quantitative evidence of how these domains relate to the functional impact of the disorder. In this preregistered study, we use adolescent self-report (n = 153) and parent-report (n = 106) responses in the depression section of the Development and Well-Being Assessment (DAWBA) to examine what information relevant to reported depression impact may be missing from its structured symptom-scale. We apply Large Language Models (LLMs) to participant free-text answers to extract information on a taxonomy of 80 outcomes. Through a sequential residual stacking framework that estimates incremental information in symptom sum-scores (*S*), item-level symptom-scale answers (*Q*), and LLM-derived text representations (*T*), we find that parent text contains information relevant to their report of depression impact that is not available in the structured symptom-scale, with the largest contributions concentrated in functioning-related domains (e.g., professional responsibilities, social exclusion, autonomy, coping). No such evidence was found in adolescent self-reports. Given the moderate sample size, these results are preliminary. Nonetheless, we provide a proof-of-concept framework for extracting information from open-ended free text in large existing datasets, and estimating incremental information across sources, which could be applied to uncover specific aspects of adolescent mental disorders across populations.

## 1 Introduction

Indicators of poor mental health among adolescents have risen in the past decades. In the United States, rates of major depressive episodes among adolescents increased by 52% (8.7% to 13.2%) from 2005-2017 (Twenge, Cooper, Joiner, Duffy, & Binau, 2019), and persistent sadness or hopelessness among U.S. high school students rose from 30% in 2013 to a peak of 42% in 2021 before falling to 40% in 2023 (Verlenden et al., 2024). Adolescent depression has also been found to be highly predictive of prolonged struggle with mental health into adulthood, with an estimated 2.78 times increased likelihood of adult depression when diagnosed as an adolescent, and additional associations with adult anxiety and suicidality (Johnson, Dupuis, Piche, Clayborne, & Colman, 2018). However, the majority of Major Depressive Disorder (MDD) research has been conducted in adults, despite numerous studies suggesting potential differences in manifestation, latent structure, and response to treatment of the disorder between adolescents and adults (Rice et al., 2019; Wight, Sepúlveda, & Aneshensel, 2004). We aim to address this gap directly through perspectives of adolescent experiences.

MDD is an inherently hard phenomenon to measure. It is heterogeneous and manifests in many different ways, with low overlap in symptoms across individuals (Fried & Nesse, 2015) and moderate overlap across established scales (Fried, 2017, 2020). A systematic review of 138 studies spanning 170 different study populations by Haroz et al. (2017) found that only seven of fifteen frequently mentioned features of depression worldwide are part of the DSM criteria for MDD. Similarly, a study of 8790 Brazilian students found numerous features that differentiated depressed and non-depressed participants, and yet were absent from the DSM and ICD (Manfro et al., 2023). Failing to adequately measure the effects of depression that people experience, and care about, contributes to incomplete assessments of severity (Stringaris & Goodman, 2013) and skewed perspectives on treatment efficacy.

In response, numerous qualitative studies have been conducted to derive factors relevant to depressed people that may not be captured adequately in standard instruments. Krause, Midgley, Edbrooke-Childs, and Wolpert (2021) conducted a post-hoc qualitative analysis of interview data collected for the IMPACT-ME study (Goodyer et al., 2011; Midgley, Ansaldo, & Target, 2014), with adolescent, parent, and therapist participants. A taxonomy of 29 outcome domains most prominently discussed by participants was derived, with very few of them consistently reported in recent quantitative treatment outcome studies. Similarly, Chevance et al. (2020) surveyed patients reporting a depressive disorder, informal care-givers, and health-care professionals about their perspectives of the expected benefits of depression treatment. From these responses, a taxonomy of 80 outcome domains was derived. These studies identify what matters to people with depression and those around them, but do not directly quantify how much these additional domains contribute to the impact a disorder has on a person’s life. Testing this quantitatively may be accomplished with: 1) an open description of experiences from participants alongside structured symptom measures, 2) a measure of the impact of depression, and 3) a method for extracting numerical representations of potentially relevant domains from those descriptions.

The Development and Well-Being Assessment (DAWBA) provides the first two requirements (R. Good-man, Ford, Richards, Gatward, & Meltzer, 2000). It features responses from adolescents, their parents and, occasionally, their teachers on a set of questionnaires, impact ratings, and structured free-text interviews for a number of mental-health disorders, including depression. Recent advancements in natural language processing (NLP), specifically in large language models (LLMs), have made the third possible. For example, Xin et al. (2026) developed logistic regression classifiers operating on text embeddings to classify transcribed interview text as exemplifying a given outcome, after training on human-coded labels (Krause et al., 2021). However, this framework of training classifiers operating on text embeddings requires large amounts of human-coded data, which may not always be available for the outcomes of interest. The latest LLMs make a different approach feasible, namely to predict the presence (or absence) of each outcome directly, after reading a prompt defining the outcome and the text to be classified. In this way, it is possible to efficiently quantify elements of already-collected semi-structured text at a large scale.

In this preregistered study (Matar, Lossio-Ventura, Pereira, & Nielson, 2026), we combine these elements to test whether free text contains information relevant to the impact of depression beyond that which is present in symptom rating scales. Using already-collected depression data from adolescent and parent responses to the DAWBA, we hypothesize that text adds information above symptom rating scale sum-scores, item-level symptom ratings, and their combination. We also hypothesize that supplementing sum-scores with information derived from text will be more predictive than when supplementing with individual symptom ratings. We test this with a residual stacking framework that allows for the isolation of contributed signal to specific sources, and additionally attribute added signal to specific outcome domains. In testing our hypotheses, we also demonstrate an analytical framework that makes it possible to exploit existing free-text data to discover and assess the importance of previously unmeasured aspects of adolescent mental health disorders across cultures, ages, and/or sex. All hypotheses were motivated by exploratory analyses in a development set. Models frozen prior to confirmatory analyses were fit on the development set and evaluated on participants in the held-out set. Hypotheses were tested separately in adolescent and parent responses from the held-out participants.

## 2 Materials & Methods

### 2.1 Data

#### 2.1.1 Collection & Pre-processing

A collection of 347 paired adolescent-parent samples was obtained between Summer 2017 and Spring 2021 for a broader longitudinal study: *the National Institute of Mental Health Characterization and Treatment of Adolescent Depression (CAT-D) study* (NIH IRB protocol number 18-M-0037, registered on clinicaltrials.gov as NCT03388606). This study is described in further detail by Gorham et al. (2022) and Sadeghi et al. (2022). Participants were screened under NIH IRB protocol 01-M-0254 prior to enrolling in CAT-D, and all participants who screened for CAT-D were considered for this analysis, regardless of their eligibility or enrollment in CAT-D. Participants were asked to respond to the Development and Well-Being Assessment (DAWBA; R. Goodman et al., 2000), with a financial incentive of $30, in addition to completing a number of other tasks. Twenty participant pairs were removed due to withdrawal of consent. Only participant meta-data and responses to the depression section of the DAWBA were considered for our analyses. Free-text responses were manually de-identified prior to inclusion in this study.

#### 2.1.2 Data Split & Further Processing

The preprocessed dataset of 327 paired parent-adolescent records was initially split into development (n=219, *∼* 67%) and held-out (n=108, *∼* 33%) sets, stratified by eligibility to the CAT-D study. The following processing steps were decided upon and performed in the development set, followed by the held-out set after preregistration. While all of these data were used as part of the screening process for the CAT-D study, we did not access the held-out set for other purposes prior to the execution of the preregistered analyses.

Each adolescent participant is the subject of both a self-report and parent-report response set. These were separated into two response samples (adolescent and parent), and all subsequent processing and analyses were conducted separately within each. Exclusion criteria were applied to responses independently, such that excluding an informant’s response in one sample did not necessarily remove the matched response in the other sample. Consequently, the final samples contain partially overlapping, rather than matched, sets of adolescent self- and parent-responses. Additionally, splitting into development and held-out sets prior to separating the two response samples ensured that there was no overlap of adolescent participants across splits.

Four symptom-scale items (18i-18l, covering ideation and attempt of self-harm/suicide) were largely missing in both samples and thus excluded from analysis. The DAWBA uses skip logic, prompting informants to complete later sections only on the basis of their earlier answers. As such, the responses of informants not prompted to answer semi-structured free-text questions were removed. Missing values for symptom-scale items as a result of not being prompted were simply marked as 0. For all other cases of missing entries in symptom-scale items, the whole response was removed. Responses with 6 or more missing text entries were dropped, and any missing answers were filled as *“Not answered”* for responses with 5 or fewer missing entries. After all cleaning steps, the adolescent and parent development sets contained 103 and 71 responses, respectively, over 120 unique adolescent participants. All processing steps described were applied to the adolescent and parent held-out sets as well, resulting in 50 and 35 responses, respectively, over 60 unique adolescent subjects. All filtering and response exclusions are outlined in Figure 1.

**Figure 1:**
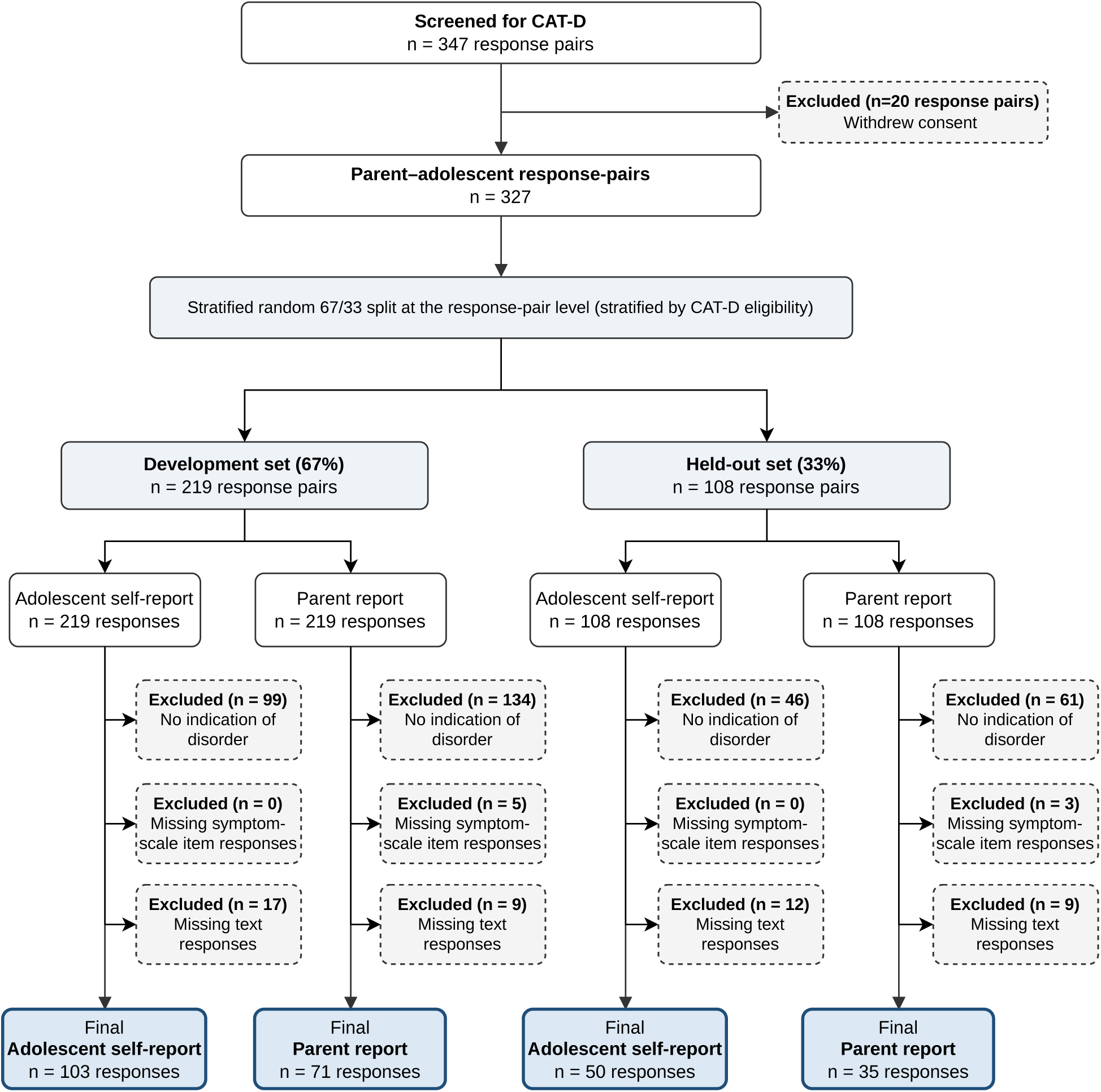
Participant flow from screening to final analysis samples.

#### 2.1.3 Sample Characteristics

Table 1 displays characteristics of the adolescent participants in the development and held-out splits for both adolescent self-report and parent report. All adolescent participants were between ages 11 and 17. The distribution of symptom sum-scores was comparable across both splits and adolescent-/parent-response samples, although with a slightly higher mean in the adolescent self-response sample. Impact sum-scores were similarly reported at comparable magnitudes across the four groups. Most adolescent participants were female (61*−*70%), and the vast majority of them were screened as having MDD (98*−*100%). The adolescent-response sample included a slightly higher proportion of participants eligible for CAT-D (80 *−* 81%) than the parent-response sample (66 *−* 70%).

**Table 1:** Participant characteristics by informant and data split. MDD abbreviates Major Depressive Disorder. HV abbreviates Healthy Volunteer.

|  | **Adolescent** |  | **Parent** |  |
|---|---|---|---|---|
| **Feature** | **Development**<br>(*n* = 103) | **Held-out**<br>(*n* = 50) | **Development**<br>(*n* = 71) | **Held-out**<br>(*n* = 35) |
| **Age, Avg** | 15.49 | 15.63 | 15.50 | 15.48 |
| Median | 15.76 | 16.07 | 15.98 | 16.00 |
| IQR | 14.44–16.53 | 14.32–16.95 | 14.17–16.55 | 14.50–16.68 |
| **Symptom sum-score, Avg** | 17.70 | 18.08 | 17.20 | 16.23 |
| Median | 18.00 | 18.00 | 17.00 | 16.00 |
| IQR | 15.00–20.00 | 16.25–20.00 | 14.00–21.00 | 13.00–20.50 |
| **Impact sum-score, Avg** | 10.73 | 11.14 | 11.03 | 11.17 |
| Median | 11.00 | 12.00 | 11.00 | 11.00 |
| IQR | 9.00–13.00 | 9.25–15.00 | 9.00–13.00 | 9.00–14.00 |
| **Sex, n (%)** |  |  |  |  |
| Female | 72 (69.9%) | 35 (70.0%) | 43 (60.6%) | 24 (68.6%) |
| Male | 31 (30.1%) | 14 (28.0%) | 28 (39.4%) | 11 (31.4%) |
| Nonbinary | 0 (0.0%) | 1 (2.0%) | 0 (0.0%) | 0 (0.0%) |
| **CAT-D Eligibility, n (%)** |  |  |  |  |
| Eligible | 84 (81.6%) | 40 (80.0%) | 50 (70.4%) | 23 (65.7%) |
| Ineligible: meets exclusion criteria | 15 (14.6%) | 7 (14.0%) | 17 (23.9%) | 12 (34.3%) |
| Ineligible: does not meet inclusion criteria | 4 (3.9%) | 3 (6.0%) | 4 (5.6%) | 0 (0.0%) |
| **Participant type, n (%)** |  |  |  |  |
| MDD | 101 (98.1%) | 49 (98.0%) | 71 (100.0%) | 35 (100.0%) |
| HV | 1 (1.0%) | 1 (2.0%) | 0 (0.0%) | 0 (0.0%) |
| Anxious | 1 (1.0%) | 0 (0.0%) | 0 (0.0%) | 0 (0.0%) |

#### 2.1.4 Response Characteristics

Table 2 displays characteristics of free-text answers of adolescents and parents in the development and held-out sets. Nearly all 10 questions (displayed in section 9.4.1 of the Supplemental Materials) were answered on average across all groups (average 9.87-9.94). Median total response length was higher in the development set than in the held-out set for both adolescents (198 vs. 163 words) and parents (249 vs. 182 words). Additionally, parents tended to write more than adolescents in both the development (249 vs. 198 words) and held-out (182 vs. 163 words) sets. Interquartile ranges overlapped substantially, however, and no formal comparisons were made, so these differences should be read as descriptive.

**Table 2:**
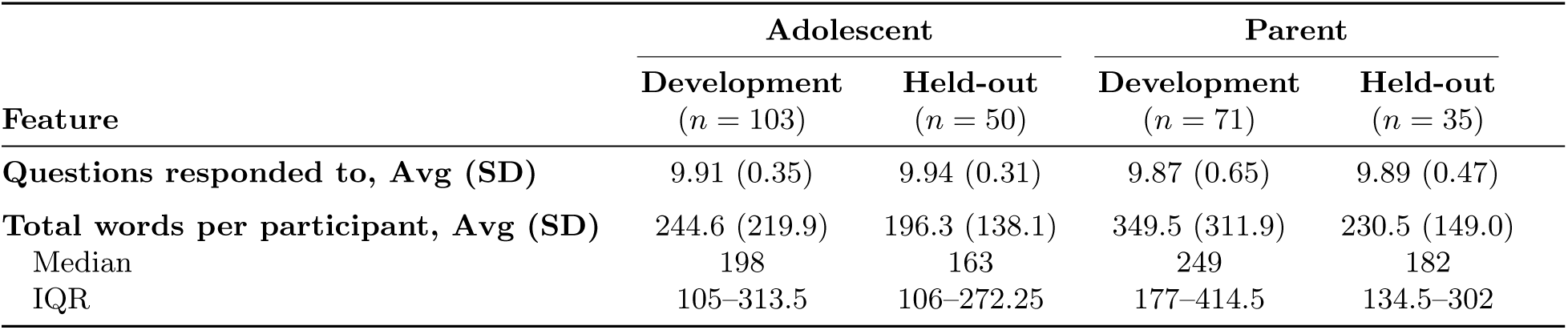
Free-text response characteristics by informant and data split. Text length refers to the number of words in the concatenated free-text answers.

|  | **Adolescent** |  | **Parent** |  |
|---|---|---|---|---|
| **Feature** | **Development**<br>(*n* = 103) | **Held-out**<br>(*n* = 50) | **Development**<br>(*n* = 71) | **Held-out**<br>(*n* = 35) |
|---|---|---|---|---|
| **Questions responded to, Avg (SD)** | 9.91 (0.35) | 9.94 (0.31) | 9.87 (0.65) | 9.89 (0.47) |
| **Total words per participant, Avg (SD)** | 244.6 (219.9) | 196.3 (138.1) | 349.5 (311.9) | 230.5 (149.0) |
| Median | 198 | 163 | 249 | 182 |
| IQR | 105–313.5 | 106–272.25 | 177–414.5 | 134.5–302 |

### 2.2 Residual Stacking Prediction Framework

The target outcome for prediction is *Impact Score*, which is taken as the sum of five depression impact questions at the end of the depression section of the DAWBA (shown in Table 12 in the Supplemental Materials). This target is predicted within a residual stacking framework, which makes it possible to isolate the incremental signal present in three classes of predictor features. The first class of features, denoted as *S*, is the sum of 23 individual items in the depression symptom-scale section of the DAWBA, with a possible discrete range of [0,25]. The second, *Q*, is a 23-dimensional vector of item-level answers to the symptom-scale section of the questionnaire, with response options of *{*0,1*}* or *{*0,1,2*}*. The third class is *T*, which is an 80-dimensional numerical representation of informant answers to free-text prompts. The signal of a given class of features is quantified as the out-of-sample coefficient of determination (*R*^2^) of a linear regression predicting *Impact Score* from the input features chosen. This metric was selected over others such as MSE, RMSE, or MAE because it provides insight into performance relative to a simple guess of the mean, rather than raw error.

In each sample, *S*, *Q*, and *T* were used to predict *Impact Score*. A number of residual stacking setups were explored, including single-layer (*S, Q, T*), two-layer (*ST, QT, SQ*), and three-layer (*SQT*) setups. All *S*-layer models were simple linear regressions, whereas *Q* - and *T*-layers were ridge regressions, regardless of position in a given stacked setup. Multi-layer setups were implemented such that a given layer’s residuals served as the target of the subsequent layer. For example, in the *SQT* model, a linear regression was fit on *S* to predict *Impact Score*, then a ridge regression model was fit on *Q* to predict the error of the *S* model. This resulted in an *SQ* model whose final output was the sum of *S*-model outputs and its predictions of *S* residuals. Finally, another ridge regression model was fit on *T* to predict the error of the *SQ* model, whose final outputs were the sum of the *SQ* model and its prediction of the *SQ* model error. This framework is outlined in Table 3.

**Table 3:**
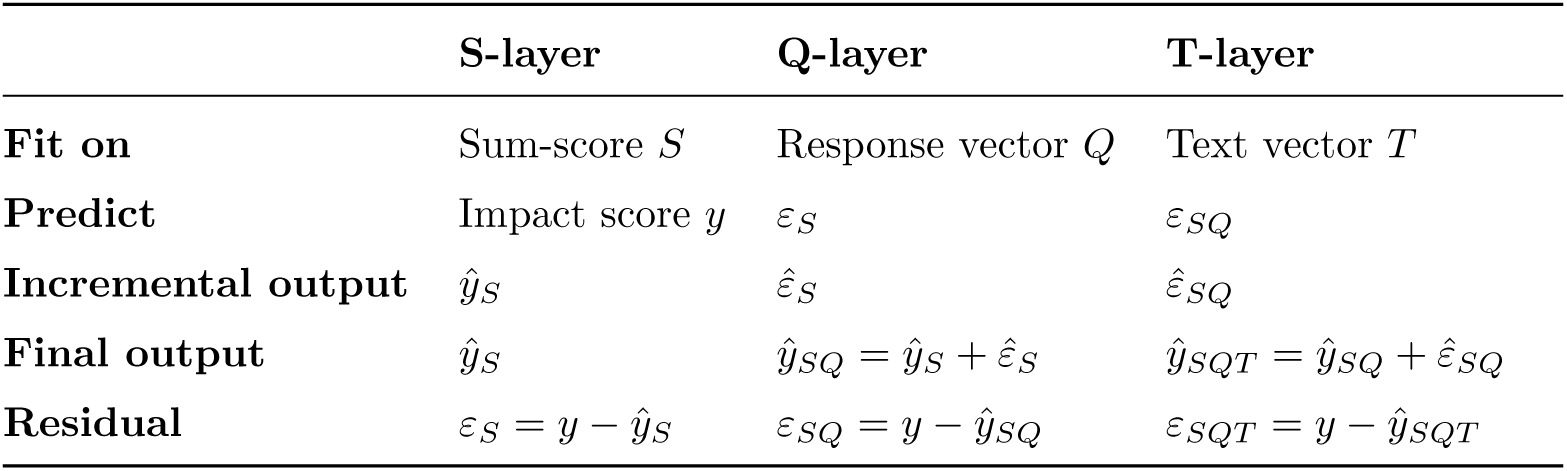
Residual stacking framework by layer for *SQT*. *ε_X_* denotes the residual after layer *X*; *ε*^*_X_* denotes its prediction by the next layer.

|  | **S-layer** | **Q-layer** | **T-layer** |
|---|---|---|---|
| **Fit on** | Sum-score *S* | Response vector *Q* | Text vector *T* |
| **Predict** | Impact score *y* | ε<sub>*S*</sub> | ε<sub>*SQ*</sub> |
| **Incremental output** | *ŷ*<sub>*S*</sub> | ε̂<sub>*S*</sub> | ε̂<sub>*SQ*</sub> |
| **Final output** | *ŷ*<sub>*S*</sub> | *ŷ*<sub>*SQ*</sub> = *ŷ*<sub>*S*</sub> + ε̂<sub>*S*</sub> | *ŷ*<sub>*SQT*</sub> = *ŷ*<sub>*SQ*</sub> + ε̂<sub>*SQ*</sub> |
| **Residual** | ε<sub>*S*</sub> = *y* − *ŷ*<sub>*S*</sub> | ε<sub>*SQ*</sub> = *y* − *ŷ*<sub>*SQ*</sub> | ε<sub>*SQT*</sub> = *y* − *ŷ*<sub>*SQT*</sub> |

The difference in *R*^2^ (variance explained) between *S* /*SQ* or *SQ* /*SQT* provides insight into incremental signal available in *Q* or *T* when they are added. Furthermore, any shared variance is assigned to the lower level models, so that the difference in out-of-sample *R*^2^ between *SQ* and *SQT* only represents the *unique* additional variance explained by *T*. We use the out-of-sample *R*^2^ because in-sample *R*^2^ is trivially non-decreasing as features are added, but out-of-sample *R*^2^ reflects generalized performance and penalizes overfitting. All the prediction models tested were fit using the entirety of the development set, and out-of-sample prediction results were obtained by deploying them on the held-out set.

#### 2.2.1 T-layer Configurations

The development set was used to select lambda regularization penalties for *Q* - and *T*-layers, as well as a number of methods and configurations to derive numerical representations of text. The first, *LR*, involved embedding text and using the resulting numeric vectors as inputs to previously trained logistic regression models for each outcome domain, similar to work done by Xin et al. (2026). The second, *LLM Prompting*, involved prompting LLMs to test the presence of the outcome domain in the text directly. The ultimate goal in both cases was to represent text as a numeric vector reflecting some taxonomy of mental health symptoms/outcomes/domains. This exploration is described in further detail in the Model Configuration Selection section of the Supplemental Materials.

Ultimately, we chose *LLM Prompting* as the method of derivation, using the taxonomy of 80 mental health outcomes derived by Chevance et al. (2020). For this method, the 10 free-text items, including both question and answer, were first concatenated into one large block of text before wrapping in a broader prompt, which gave context to the text and provided instructions for what to output. LLMs were prompted independently for each outcome in the taxonomy and for each response. The resulting feature vector for each instance of *T* was an 80-dimensional concatenation of LLM output.

There were multiple decisions to make regarding the configuration of LLM prompting, which we will refer to as *pseudo-hyperparameters*. Namely, these involve the LLM base-model, temperature, prompting method, and output type. Pseudo-hyperparameters were selected separately for each residual setup including *T* (*T, ST, QT, SQT*) within each response-sample, for a total of eight selected sets of pseudo-hyperparameters. LLM base-models were selected from *Gemini 2.5 Pro, Qwen 3.6 27B* and *Llama 3.3 70B Instruct*. *Qwen* and *Llama* are open-weight models, which we ran on local hardware. *Gemini* was run through HHS Enterprise Gemini with FedRAMP High authorization. These models, developed by different entities, differ in size, capability, and price. Estimated performance rankings varied across residual setups and response-samples in the development set. Temperature, a true hyperparameter of LLMs, controls the stochasticity of output. Smaller temperature settings result in more deterministic and consistent output across runs, but may limit variation in LLM output and ability to capture nuance. We also considered two different LLM prompting approaches. In our *zero-shot* prompting, we provided a description of the source of the text, a concatenated block of DAWBA free-text questions and answers, the target domain, a definition of that domain, and instructions for output. In contrast, our *few-shot* prompting supplemented the provided description of the domain with a few examples of text and expected output, providing further information about the intended interpretation of the domain. An important consideration of *few-shot* prompting is that, if provided examples are too narrow, LLMs may not be able to generalize or capture more nuanced information as intended. Similarly, if they are too broad, LLMs may have a wider view of a domain than is appropriate. Further, all chosen pseudo-hyperparameter sets employed a *Scale* output type, which prompted LLMs to output the severity (*{*0,1,2,3,4*}*) of an outcome. This is in contrast to the *Binary* output type, which prompted LLMs to simply output whether the outcome was present in the text or not (*{*0,1*}*). An example of *zero-shot, scale* prompting is displayed in the Supplemental Materials.

These pseudo-hyperparameters were selected alongside lambda (*λ*), a true regularization hyperparameter of ridge regression. Generally, larger values of lambda place higher penalty on learned coefficients and constrain the model fit to the training set, reducing the likelihood of over-fitting. These values in *Q* - and *T*-layers, alongside pseudo-hyperparameters relevant only to *T*-layers, were selected during exploratory analyses within the development set, and are shown in Table 4. The full explored search space of pseudo-hyperparameters, in addition to selection criteria, are outlined in the Supplemental Materials.

**Table 4:** Final frozen model configurations selected during exploratory analysis using the development set, by model set-up and informant. All selected configurations use the *Scale* output type. Model families are Llama 3.3 70B Instruct, Qwen 3.6 27B, and Gemini 2.5 Pro; *few* and *zero* denote few-shot and zero-shot prompting, respectively. *λ_Q_* and *λ_T_* denote ridge regularization for the *Q*-and *T*-layer regressions, respectively.

| **Set-up** | **Informant** | **LLM<br>Model** | **LLM<br>Prompt** | ***λ<sub>Q</sub>*** | ***λ<sub>T</sub>*** | **Temp.** |
|---|---|---|---|---|---|---|
| *S* | Parent | – | – | – | – | – |
|  | Adolescent | – | – | – | – | – |
| *Q* | Parent | – | – | 5 | – | – |
|  | Adolescent | – | – | 15 | – | – |
| *T* | Parent | Llama | few | – | 5 | 1.0 |
|  | Adolescent | Gemini | few | – | 5 | 0.4 |
| *SQ* | Parent | – | – | 15 | – | – |
|  | Adolescent | – | – | 50 | – | – |
| *ST* | Parent | Llama | few | – | 15 | 0.0 |
|  | Adolescent | Gemini | few | – | 25 | 0.4 |
| *QT* | Parent | Qwen | zero | 5 | 50 | 0.0 |
|  | Adolescent | Gemini | few | 15 | 5 | 0.4 |
| *SQT* | Parent | Qwen | zero | 15 | 75 | 0.0 |
|  | Adolescent | Gemini | few | 50 | 15 | 0.4 |

### 2.3 Statistical Analyses

Our primary goal was to determine if there was information relevant to the impact of depression in the free-text responses, beyond that captured by the symptom rating scale. This was operationalized through two separate sets of hypotheses, each tested independently in adolescent- and parent-response samples through a residual stacking prediction framework. These hypotheses were determined and preregistered prior to conducting confirmatory analyses (Matar et al., 2026).

#### 2.3.1 Hypotheses 1A/1B

The first set of hypotheses assessed if free text (*T*) carries signal in predicting depression impact incremental to what is found in granular symptom-scale items (*Q*) and their sum-scores (*S*). This can be represented formally as:

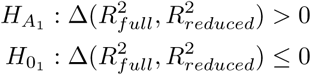

where Δ(*x, y*) represents the difference *x − y*, 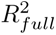 is the *R*^2^ in the held-out set of some full model including *T*, and 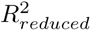 represents that of some reduced model excluding *T*. Three full/reduced pairs were evaluated, resulting in three sub-hypotheses: (*ST*, *S*), (*QT*, *Q*), and (*SQT*, *SQ*), with a single p-value derived for each pair. This was performed separately for each of the adolescent- and parent-response samples, corresponding to Hypotheses 1A and 1B, respectively.

The null hypothesis states that *T* carries no predictive information, meaning that a model including *T* performs no better than the same model without it. To construct this null, a 1000-iteration permutation procedure was employed. In each iteration, entire text outcome severity vectors (*T*) in the development set were shuffled according to a unique random seed, stratified by CAT-D eligibility group. The original prediction target variable (impact score) and reduced-model sources (*S*, *Q*, *SQ*) were kept fixed. This breaks the connection between the LLM-derived vectors and the prediction target, while maintaining any correlation between the presence of outcomes within responses. The same shuffle was used for all three full models within each iteration—each of *ST*, *QT*, and *SQT* was refit on the shuffled development set and evaluated on the un-shuffled held-out set, yielding a null *R*^2^ for each setup in each iteration. The reduced models (*S*, *Q*, *SQ*) contained no text features and were therefore unaffected by the shuffle; their observed held-out set *R*^2^ values were held fixed across iterations, so each iteration yielded three null differences 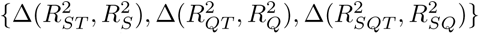. P-values were derived via the Westfall-Young step-down minP procedure (Westfall & Young, 1993) to control family-wise error rate (FWER) at *α* = 0.05, which is described in further detail in the Supplemental Materials.

#### 2.3.2 Hypotheses 2A/2B

Second, we assessed if the incremental signal of free text (*T*), as a supplement to symptom sum-scores (*S*), exceeds that of granular symptom-scale question items (*Q*). This can be represented formally as:

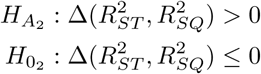

where Δ(*x, y*) represents the difference *x − y*, and 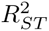 and 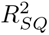 represent the held-out set *R*^2^ of the stacked *ST* and *SQ* regression models, respectively. Similar to Hypotheses 1A/1B, Hypothesis 2A refers to a separate test of the adolescent-response sample, whereas Hypothesis 2B refers to that of the parent-response sample.

P-values were again calculated non-parametrically, deriving a null distribution by randomly swapping held-out set predictions from *ST* and *SQ* over 1000 permutations, simulating exchangeability of *SQ* and *ST*. P-values were calculated as the proportion of null differences as or more extreme than what was observed:

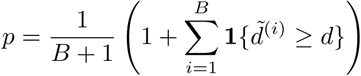

where *B* = 1000 is the number of permutation iterations, *d* is the observed difference 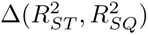, and *d̃*^(*i*)^ is the null difference in permutation iteration *i*. No correction was employed, and significance was set at *p <* 0.05.

#### Deviation from Preregistration

An adaptation of the Westfall and Young (1993) step-down maxT procedure was preregistered for Hypotheses 1A/1B, using raw *R*^2^ differences as test statistics instead of normalized t-statistics. However, raw differences in *R*^2^ are not pivotal, as 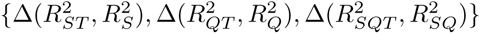 are on differing scales. Thus, a minP approach was employed instead, since p-values are pivotal and invariant to scale. Results from the minP approach are used to draw conclusions, and adapted maxT results are reported for completeness.

## 3 Results

Model configurations, including pseudo-hyperparameters and lambda regularization values, were selected in the development set and frozen prior to confirmatory testing. The entire development set was used to train models according to frozen configurations. Trained models were subsequently evaluated on the held-out set. Note that differences and relative changes were computed from unrounded values and may therefore differ slightly from differences between the rounded values reported.

### 3.1 Prediction performance of each setup

For insight into standalone predictive ability, each of *S, Q, T, SQ, ST, QT,* and *SQT* residual stacking setups were evaluated in both response samples. The performance of each residual setup for adolescent and parent samples is displayed in Table 5, along with 95% confidence intervals (CIs) derived via 1000-iteration bootstrapping.

**Table 5:** Held-out set *R*^2^ (95% CIs) performance of models by residual setup and response-sample. Models were fit on the development set only. Values are rounded to the nearest thousandth.

|  | Response sample |  |
|---|---|---|
| Residual setup | PARENT | ADOLESCENT |
| S | 0.297 (−0.074, 0.491) | 0.144 (−0.076, 0.277) |
| Q | 0.272 (−0.213, 0.541) | 0.240 (0.029, 0.348) |
| T | 0.073 (−0.370, 0.342) | −0.071 (−0.443, 0.159) |
| SQ | 0.341 (−0.043, 0.568) | 0.220 (0.012, 0.348) |
| ST | 0.336 (−0.041, 0.555) | 0.193 (−0.036, 0.349) |
| QT | 0.340 (−0.114, 0.604) | 0.133 (−0.147, 0.305) |
| SQT | 0.400 (0.035, 0.623) | 0.221 (−0.007, 0.367) |

Held-out set performance for *S* was higher than *Q* in the parent sample but lower in the adolescent sample, with *T* consistently performing the worst of the single-source setups. Held-out performance in the parent sample was comparable across all 2-layer setups (*R*^2^ *≈* 0.34), with a slightly better performance in the 3-layer *SQT* setup (*R*^2^ = 0.400). In the adolescent-response sample, *SQ* and *SQT* performed comparably (*R*^2^ *≈* 0.22) with slightly worse performance in *ST* (*R*^2^ = 0.193) and *QT* (*R*^2^ = 0.133), although CIs overlapped heavily. Note that out-of-sample *R*^2^ values can be negative, although this only happened for one model. We also carried out cross-validation experiments within the development set to make design choices, and to select the models that we applied to the held-out set. Results of these experiments are summarized in Table 8 in the Supplemental Materials.

### 3.2 Incremental prediction value of text-derived features

Table 6 summarizes the incremental change in *R*^2^ when predicting depression impact in the held-out set (Δ*R*^2^), obtained by adding outcome severity features derived from text. The experiments were carried out against every possible baseline model (S, Q, SQ), in a residual stacking framework, over parent and adolescent samples. The table also reports 95% CIs derived via 1000-iteration paired bootstrapping, and the respective p-values. Significance was set at *p <* 0.05. Cross-validated results within the development set are available in Table 8 in the Supplemental Materials.

**Table 6:** Differences (95% CIs) in held-out set *R*^2^ performance of models predicting depression impact in parent-and adolescent-report samples, computed from unrounded values. All models fit on the development set only. Values are rounded to the nearest thousandth. FWER-adjusted p-values from the Westfall–Young step-down minP (*p*_minP_) and maxT (*p*_maxT_) procedures are reported for *SQT/SQ, ST/S,* and *QT/Q*. Raw unadjusted p-values are reported for *ST/SQ*. Bold and * denote *p <* 0.05.

| Model<br>comparison | PARENT |  |  | ADOLESCENT |  |  |
|---|---|---|---|---|---|---|
|  | Δ*R*<sup>2</sup> | *p*<sub>minP</sub> | *p*<sub>maxT</sub> | Δ*R*<sup>2</sup> | *p*<sub>minP</sub> | *p*<sub>maxT</sub> |
| SQT − SQ | 0.060 (0.002, 0.121) | **0.033**\* | 0.059 | 0.001 (−0.117, 0.110) | 0.49 | 0.43 |
| QT − Q | 0.068 (0.003, 0.139) | **0.033**\* | **0.043**\* | −0.107 (−0.299, 0.059) | 0.76 | 0.76 |
| ST − S | 0.039 (−0.112, 0.180) | 0.11 | 0.11 | 0.049 (−0.051, 0.142) | 0.26 | 0.22 |
|  | Δ*R*<sup>2</sup> | *p*<sub>unadj</sub> |  | Δ*R*<sup>2</sup> | *p*<sub>unadj</sub> |  |
| ST − SQ | −0.004 (−0.268, 0.247) | 0.67 | – | −0.027 (−0.154, 0.090) | 0.48 | – |

#### Hypotheses 1A/1B

Differences in variance explained between full and reduced model pairs for Hypothesis 1A (adolescent) in the held-out set ranged from *−*0.107 to 0.049, with no significant differences, whereas increments ranged from 0.039 to 0.068 for Hypothesis 1B (parent), resulting in two significant findings: 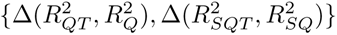. *R*^2^ increased by 25% with the addition of *T* over *Q*, resulting in an explained variance increment of 0.068. This was 9% of the remaining variance unexplained by *Q* and accounted for 20% of the total variance explained by *QT* (0.340), ultimately resulting in a significant corrected p-value of 0.033. Similarly, *R*^2^ increased by 17% from *SQ* (*R*^2^ = 0.341) to *SQT* (*R*^2^ = 0.400) in the held-out parent sample, with a raw effect size of 0.060. The addition of *T* accounted for 9% of the remaining variance unexplained by *SQ*, and corresponds to 15% of the total variance explained by *SQT*. The stepwise corrected p-value for this sub-hypothesis was also *p* = 0.033. In all other cases, including all three pairs for Hypothesis 1A (adolescent-report) and 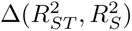 in Hypothesis 1B (parent-report), corrected p-values exceeded 0.05, and we could not reject the null hypothesis. Additionally, all 95% CI estimates, which describe the sampling uncertainty, contained 0 for these cases.

The failure to reject the null for the difference between *ST* and *S* in the parent sample was notable because the null was rejected for *QT* and *SQT*. Additionally, *ST* was the only model of the three built on Llama instead of Qwen. As a post-hoc sensitivity analysis, we additionally tested the difference between *ST* and *S* with the best performing Qwen model from the development set (full results with multiple comparisons corrected for this additional comparison are in Table 11 in the Supplemental Materials). With the Qwen model, *ST* explained significantly more variance than *S* after multiple comparisons correction (Δ*R*^2^ = 0.088, *p* = 0.04), and the other comparisons maintained their significance.

#### Hypotheses 2A/2B

For 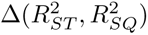, observed increments are *−*0.004 and *−*0.027 for parent and adolescent response samples, respectively, and 95% CIs contained 0 for both. No p-value was less than the pre-determined threshold of 0.05, thus we could not reject the null hypothesis for 2A or 2B.

### 3.3 Exploratory Feature Contribution Analysis

In addition to assessing if incremental signal exists in free text, we aimed to attribute this signal to specific symptomatic and functional domains to determine how informative, on average, each given feature is. Using LMG-decomposition (Lindeman, Merenda, & Gold, 1980), *R*^2^ contributions of *T* were apportioned to each of the 80 outcomes that make up each free text’s numerical representation vector. The attributed signal of each feature is the average increase in *R*^2^ when it is sequentially added to a model, averaged over 2000 random feature orderings. This was also computed in the development set using out-of-fold (OOF) predictions averaged over each of 30 folds from a 10-time repeated 3-fold cross-validation (CV) framework.

Both development and held-out set feature analyses are considered exploratory and hypothesis generating, rather than confirmatory. A subset of the 80 outcomes is displayed in Figure 2, chosen as the union of the top 10 contributors (those with the largest average *R*^2^ contribution) in each of the development/held-out *QT* /*SQT* parent-response models, as these were the only response-sample/configuration combinations to survive FWER corrections in favor of their corresponding alternative hypothesis (1B).

**Figure 2:**
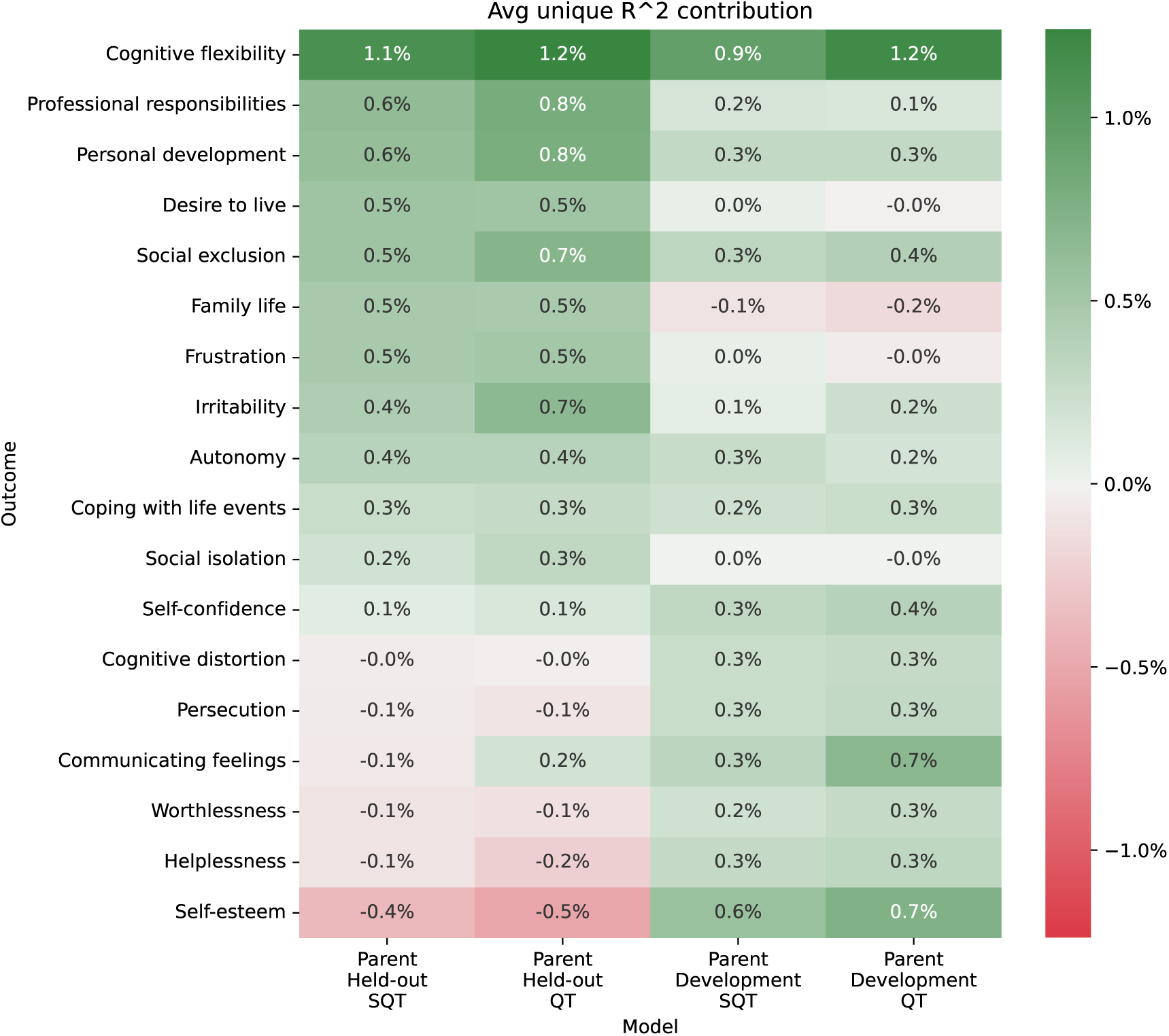
Aggregated increment in % variance explained (*R*^2^) when an outcome is added to the model (one of *QT* or *SQT* in the parent-response sample). Displayed values are mean out-of-sample performance increments across 2000 orderings in the held-out set, and average out-of-fold performance increments across 2000 orderings and 10-time repeated 3-fold CV (30 folds) in the development set.

Across all four response-sample/configuration combinations, the *Cognitive flexibility* outcome consistently provided the highest average contribution to variance explained (0.9 *−* 1.2%), followed by *Professional responsibilities* (0.1 *−* 0.8%), *Personal development* (0.3 *−* 0.8%), *Social exclusion* (0.3 *−* 0.7%), *Irritability* (0.1 *−* 0.7%), *Autonomy* (0.2 *−* 0.4%), *Coping with life events* (0.2 *−* 0.3%), and *Self-confidence* (0.1 *−* 0.4%). Some outcomes, however, showed consistent positive contribution across *SQT* and *QT* in the held-out set, but detrimental or zero effect in the development set for both stacked setups. For example, *Family life* had an average contribution of 0.5% in both *SQT* and *QT* in the held-out set, but *R*^2^ was 0.1% and 0.2% lower on average in the same model set-ups in the development set, respectively. On the other hand, *Self-esteem* provided strong gains on average in the development set for *SQT* and *QT* (0.6% and 0.7%, respectively), but performance was lower on average in the held-out set (*−*0.4% and *−*0.5%, respectively). Figure 3 in the Supplemental Materials shows these contributions as proportions of the total incremental variance explained by *T*.

## 4 Discussion

The starting point for our work was the availability of the DAWBA instrument, used in the context of depression. There, adolescents and parents answer a structured questionnaire composed of symptom-scale items (*Q*), which can be aggregated into sum-scores (*S*), as well as providing short free-text narratives. Using LLMs, the latter can be transformed into quantitative features representing a taxonomy of 80 mental-health outcomes (Chevance et al., 2020) (*T*). Our main objective was to determine if there was information in free text relevant to the impact of depression that was not captured by the structured symptom-scale. We operationalized this through a residual stacking framework capable of isolating predictive capability of each source in prediction of adolescent- and parent-reported impact of depression.

We hypothesized that *T* would increase variance explained over *S*, *Q*, and stacked *SQ* models in both adolescent- and parent-response samples. This was supported in the parent-response sample over *Q* (Δ*R*^2^ = 0.068, *p_minP_* = 0.033) and *SQ* (Δ*R*^2^ = 0.060, *p_minP_* = 0.033), but not *S* (Δ*R*^2^ = 0.039, *p_minP_* = 0.11). No significant increases from *T* were found in adolescent responses (Δ*R*^2^ between *−*0.107 and 0.049, *p_minP_* between 0.26 and 0.76). We additionally hypothesized that the incremental signal of *T* as a supplement to *S* would exceed that of *Q*, but found no significant results in either sample, with near-zero or negative observed differences in performance between *ST* and *SQ* (Δ*R*^2^ = *−*0.027 and *−*0.004 for adolescent and parent responses, respectively).

Given the limited power of this proof-of-concept study, we did not aim to draw conclusions about the difference between adolescent self-report and parent-report. While we did observe a difference in significance for Hypothesis 1, we did not test the difference in incremental *R*^2^ between parents and adolescents. This difference may partly reflect the longer written answers by parents, whose median free-text word count exceeded that of adolescents by 19 in the held-out set (182 vs 163 words) and 51 in the development set (249 vs 198 words). It is also possible that there is greater inter-individual consistency between parent free-text responses than there is between adolescent responses.

Our results in the parent-response sample are consistent with semi-structured text containing information about depression impact that is not available in structured symptom-scale items. Although *T* alone explained far less variance in parent-reported impact than *S* or *Q* alone (0.073 vs. 0.297 and 0.272), its contribution was almost entirely complementary: *T* increased explained variance by 0.068 over *Q* and 0.060 over *SQ*, nearly its full standalone value. This suggests that the information *T* carries about parent-reported impact overlaps little with that captured by the symptom-scale items. This was not the case for *S* (Δ*R*^2^ = 0.039, *p_minP_* = 0.11). It is noteworthy that the two significant comparisons both used zero-shot Qwen, whereas few-shot Llama was employed for *ST*. We selected Llama for the *S* to *ST* comparison because of its superior performance in the development set. In an exploratory sensitivity analysis, *ST* did explain significantly more variance than *S* (Δ*R*^2^ = 0.088, *p* = 0.04) when free-text features were extracted with zero-shot Qwen instead of few-shot Llama. Thus, the failure to confirm this hypothesis was likely due to noise in the selection of models in the development set.

Given the indication that *T* contains predictive information disjoint from that in the symptom-scale in parent responses, it is worth exploring the nature of that unique signal. *T* was extracted from semi-structured text following a taxonomy of 80 domains originally derived from a qualitative study by Chevance et al. (2020). These outcomes were segmented into two classes of symptom-(64 total) and functioning-related (16 total) domains, as psychiatric classifications often distinguish between the two. Symptom-related domains, pertaining to the manifestations of a disorder, are among the most studied, whereas functioning-related domains, pertaining to the extent to which those manifestations affect various aspects of daily life, receive less attention (Krause et al., 2021). The two, which are both typically required for a diagnosis of MDD (American Psychiatric Association, 2013), are related but do not fully overlap. Functional impairment has been found to be only partially explained by symptom severity and, in children and adolescents, functional impact has been shown to predict later psychiatric disorder over and above baseline symptoms (Stringaris & Goodman, 2013). Thus, it is plausible to conclude that consideration of both may provide a better picture of a disorder than either alone. Because the structured DAWBA items primarily capture symptoms (R. Goodman et al., 2000), remaining relevant information not already captured might therefore be expected to concentrate in functioning-related domains.

Consistent with this, functioning-related domains were over-represented among the features contributing to *T*’s increment in the parent *QT* and *SQT* models, the only two residual setups where *T* added significant gains. Of 18 LLM-derived features, selected as the union of the top 10 contributors across development and held-out sets in each model, eight contributed positively in all four groups. Five of the eight were functioning-related domains (Professional responsibilities, Personal development, Social exclusion, Autonomy, Coping with life events), whereas three were symptom-related (Cognitive flexibility, Irritability, Self-confidence), despite functioning domains making up only 16 of the 80 in the taxonomy. Their prevalence in the original study deriving them (Chevance et al., 2020) is mixed. Professional responsibilities (11%) was among the most mentioned by all participants. Self-confidence (4.2%) was less mentioned, followed by Autonomy (4%), Coping with life events (4%), Irritability (3%), Personal development (1%), Social exclusion (*<* 1%), and Cognitive flexibility, which was only mentioned in the French cohort (*<* 1%). Suicidal Ideation (symptom-related, 23%) was the most frequently mentioned domain, but was not determined to be a major contributor when supplementing *Q* and *SQ* in our data. The predominance of functioning-related domains among these contributors may explain the discrepancy between significant results in the two responder samples, as parents tend to focus more on functioning than adolescents (Krause et al., 2021).

There are two potential interpretations relating to the prevalence of functioning-related domains in the extracted set of top contributors. The first offers that *T* recovers clinically meaningful content (the consequences of depression for school, peer, family life, etc.) that symptom-oriented structured items do not elicit. The second understands *T* as simply providing a finer-grained description of the outcome itself. The DAWBA impact score is defined as the sum of five prompts relating to distress and interference in everyday domains such as family life, friendships, and schoolwork, so parent descriptions of social exclusion or difficulties with schoolwork overlap in content with the construct being predicted. These interpretations are not necessarily mutually exclusive. A deeper understanding of these results would benefit from further analyses distinguishing between text serving to effectively extend the structured screening and text merely refining the measure of impairment that structured impact items already target.

Outside of the results’ implications in depression screenings, this study also serves as a proof-of-concept framework for evaluating the utility and/or nature of semi-structured text in relation to associated structured data, allowing for analysis of already-collected data at large scales in mental health domains. Previous work has explored similar residual stacking approaches in bioinformatics (Boulesteix & Hothorn, 2010; Boulesteix & Sauerbrei, 2011), and others have employed both feature extraction from social media postings via NLP and residual stacking broadly applied in health, psychology, and economics (Zamani & Schwartz, 2017; Zamani, Schwartz, Lynn, Giorgi, & Balasubramanian, 2018). However, to our knowledge, this is the first preregistered test of whether LLM-derived text features add predictive value above structured symptom scale questions in relation to depression impact. Although our framework is not specific to the DAWBA, the assessment is a particularly well-suited instrument for further investigations, as it was developed to be administered by lay-interviewers (R. Goodman et al., 2000), lowering barriers for administration and making collection more accessible. The DAWBA also features various skip rules throughout, making assumptions that a section is unlikely to be relevant given previous responses, and in turn reducing the burden of completion for participants. In addition to further investigations exploring the questions posed by this study, any number of taxonomies may be used to assess differences in information found in free-text across age, cultural, or sex groups, in prediction of reported impact of depression, or any other mental-health disorder in the DAWBA. There have already been numerous collections of the DAWBA across cultural (Alyahri & Goodman, 2006; A. Goodman et al., 2012; Zeinoun et al., 2013), age, and sex groups (Baser, Ford, Delgado, O’Logbon, & Cross, 2023).

### 4.1 Limitations

A major limitation of this study is the relatively small sample sizes. In the held-out set, this lowers power and decreases reliability of observed effect sizes, as evidenced by wide CIs. Model and hyperparameter selection within the development set are also affected by this, given potential variance in results, although comparisons were conducted across aggregated 10-time repeated 3-fold CV scores to minimize the volatility of model performance estimates and rankings.

Additionally, the sample was purposefully filtered to exclude participants not prompted to respond to free-text questions, removing a vast majority of healthy volunteers with 0 impact and avoiding spurious inflation of variance explained. This, paired with included participants being help-seeking, may have reduced the proportion of explainable variance by increasing the homogeneity of our sample. This may explain the low magnitude of observed *R*^2^, though these values are comparable with similar analyses predicting depression impact from symptoms (Stringaris & Goodman, 2013).

Another potential limitation is that explained variance estimates may be inflated by shared-informant variance (also referred to as shared-rater or shared-method variance), which may produce artifactual covariance between predictors (*S*, *Q*, *T*, and their stacked setups) and reported impact (Podsakoff, MacKenzie, Lee, & Podsakoff, 2003). Because the same respondent provides the symptom-scale ratings, free text, and impact ratings within each sample, variance explained may reflect genuine capture of contributors to impact, but it could also reflect the consistency of a respondent in describing the adolescent’s experiences across all three sources. Cross-informant analyses, such as predicting adolescent-reported impact from parent text, or prediction of clinician-rated outcomes, would help separate these possibilities. Additionally, statistical corrections may be effective, but likely require larger datasets than available here (Podsakoff et al., 2003).

Finally, our selected LLMs may not remain accessible for validation in the future. Gemini is closed-source and the specific version used here may be discontinued later. Open-weight models were purposefully explored with this in mind, although they present a barrier of their own by requiring access to one or more GPUs to run locally. This may also be the only option if cloud-based models cannot be used with data containing PHI or PII.

### 4.2 Future Directions

These findings, although preliminary, have implications in our understanding of adolescent depression, specifically in the operationalization of that understanding through screenings. Significant increases in parent-reported impact variance explained with the inclusion of LLM-derived text features over structured item responses suggest that semi-structured interview responses may contain explicit, unique information un-available in the structured symptom-scale.

This claim would benefit from further testing on larger samples to decrease uncertainty in observed effect sizes, increase efficacy of statistical corrections for potential shared-informant variance, and allow for rigorous cross-respondent prediction. Further, more data may uncover additional informative domains not found to be prominent in this study, and sensitivity analyses assigning uncertainty to domain contribution estimates would also allow for more definitive indications of what symptom-scales might be missing. Moreover, pairing these methods with separation of symptom- and functioning-related domains extracted from text may also provide further insight into the nature of unique information in text relevant to impact.

This project also serves as a proof of concept for a framework that has many applications. With more data, modeling paradigms more complex than regression (such as tree-based methods or neural networks) could capture non-linear feature interactions in both *Q* and *T*, increasing understanding of the relationships of factors both within and across these sources. Different taxonomies and/or combinations of taxonomies could also be explored to find differences between populations (sex, age, culture, etc) at a mass-scale, given the availability of structured and unstructured data across these groups. Additionally, beyond specifically *S*, *Q*, and *T*, this residual stacking framework may also be applied to any combination of sources, aiding in isolating signal within frameworks previously thought to be inseparable.

Finally, it may also be promising to shift the use of LLMs from deriving a score for each outcome within a taxonomy to exploring the addition of new symptom scale questions. Within this adjacent framework, participant answers to novel survey questions may be simulated and filled-in by LLMs based on free text. Prompts may be designed to capture specific aspects of mental-health, providing a proxy to test its relevance to some domain or disorder through observed differences in predictive performance when it is added as a source. As opposed to characterizing the nature of relevant information available in various sources of data, this proposed method differs from the framework described in this study in that it allows for the direct and explicit experimentation of potential changes to established screening measures, which may extend beyond the DAWBA. Allowing for the consideration of both theory and observed results in prior research to develop new structured questionnaire items, this method could leverage existing data to inform real, practical change, without the necessity of conducting new trials.

## 5 Conclusion

Depression is common and increasingly prevalent among adolescents, yet it remains less studied in this population than in adults, and is difficult to measure with structured instruments alone. Free-text descriptions from adolescents and their parents offer a way to capture aspects of the experience that fixed questionnaire items may miss. In this preregistered study, we tested whether LLM-derived features of free text add predictive signal for depression impact beyond structured symptom-scale items in the DAWBA depression section.

In parent reports, text features explained additional variance in impact beyond item-level symptom ratings, both alone (*QT* vs. *Q*) and combined with sum-scores (*SQT* vs. *SQ*). This contribution was approximately additive to that of the structured items, suggesting that parent free text carries information about impact that the symptom-scale does not. These effects were not observed in adolescent self-reports, and text did not outperform item-level symptom ratings as a supplement to sum-scores in either response sample. Exploratory attribution analyses indicated that the added variance explained by text in these parent-report models was concentrated in functioning-related domains, such as social exclusion, professional responsibilities, and autonomy, rather than in the symptoms the structured symptom-scale items are designed to assess. Whether this reflects capture of the content that the structured symptom-scale items miss or a finer-grained description of the impairment being predicted remains to be determined. Given the modest sample and wide uncertainty in effect sizes, these findings should be regarded as preliminary. More broadly, the residual stacking framework outlined here offers a way to quantify what semi-structured text adds to structured measurement using data that has already been collected. It can be extended to other taxonomies, populations, informants, and outcomes to test what any one source of data contributes to a given measure beyond the others.

## 6 Data Availability

Partial data used for these analyses are available at https://github.com/transatlantic-comppsych/CATD-study. We are only able to share data from participants who enrolled in the CAT-D study. Data from participants who only screened is not shared, but is available upon request. All code used to run analyses and produce figures is available at https://github.com/NIMH-MLT/DAWBA. Additionally, the preregistered analysis is available at https://osf.io/fs6bg.

## 7 Acknowledgments

This work utilized the computational resources of the NIH HPC Biowulf cluster (https://hpc.nih.gov).

## 9 Supplemental Materials

### 9.1 LLM-derived Numerical Representations of Text

A number of methods and configurations to derive numerical representations of text were explored in the development set, with two main classes: previously trained logistic regression (LR) and LLM prompting. The goal of both was to represent text as a vector of variables, quantifying the degree of presence in the text of items from a taxonomy of mental health symptoms/outcomes/domains.

#### Logistic Regression (LR)

Xin et al. (2026) explored the efficacy of LLMs in annotator classification of 31 mental health outcomes, taken from an original taxonomy of 30 outcomes (reported as 29 in text, but listed as 30 in their table), in transcribed interview text (Krause et al., 2021; Midgley et al., 2014). The 31 outcomes were derived from the original 30 by splitting “Other peer relationships” into “Peer relationships” and “Romantic relationships.” We similarly classified the presence of these outcomes for each of monologue (interviewee utterances) and dialogue (interviewer-interviewee) transcription segmentation types, using logistic regression classifiers trained to predict human-coded labels from embeddings of segmentations derived via Llama 3.3 70B Instruct (temperature=0.6). Due to outcome prevalence imbalances, classifiers were not developed for three of the 31 outcomes. Logistic regression hyper-parameters (C regularization factor) were derived via 4-fold CV, separately for each segmentation-type/outcome pair. The set of 56 trained classifiers (one for each of 28 outcomes for each of the two segmentation types) were then available for applying to data derived from the DAWBA.

A number of aggregation methods for the 10 question-answer pairs of DAWBA free-text was also explored: *Max, Avg, Block*. For *Max* and *Avg*, each of the 10 question-answer pairs were embedded and passed through logistic regression models separately, resulting in 10 feature vectors of length 28. The maximum and average was then taken over the 10 vectors at each position for *Max* and *Avg*, respectively, resulting in a single 28-dimensional feature vector for each. In the *Block* method, all 10 question-answer pairs were concatenated to form one large block of text, which was embedded and passed through logistic regression classifiers as a single entity, resulting in a single 28-dimensional feature vector.

Llama 3.3 70B Instruct with temperature of 0.6 was the only configuration utilized for all embeddings of this method. The class of segmentation type and aggregation method were deemed pseudo-hyperparameters, and their selection is described in further detail in Model Configuration Selection.

#### LLM Prompting

The other class of LLM-derived numerical representations involves prompting LLMs about the presence and/or intensity of some taxonomy of mental health outcomes in some input text. This was explored in two iterations. The first considered the original taxonomy of 31 outcomes (*Original*) explored in the Logistic Regression (LR) class, along with its combination with a larger taxonomy of 80 outcomes derived by Chevance et al. (2020), totaling 111 outcomes (*Combined*). The second iteration only considered the taxonomy of 80 outcomes (*Expanded*). For both iterations, the 10 question-answer pairs of free-text were concatenated using the LR *Block* method and wrapped in a broader prompt giving context to the text and instructions for what to output. Each outcome for a given taxonomy was prompted separately, and the resulting feature vector was a concatenation of LLM output of the same length as the corresponding taxonomy.

In the first iteration, the LLM was only asked to produce a binary 0/1 for the presence of a given outcome. However, two output types were explored. The *Probability* output type derived the internal token probability of ‘1’ as the output for a given outcome, whereas the *Binary* output type used the raw ‘0’/‘1’ output of the model. Additionally, iteration 1 only utilized Llama 3.3 70B Instruct with temperature 0.6.

The second iteration also explored *Binary* output, but replaced *Probability* with *Scale*, which asked for a rating of the intensity of an outcome with discrete range [0,4]. This iteration also explored Gemini 2.5 Pro and Qwen 3.6 27B in addition to Llama 3.3 70B Instruct. Further, in contrast to iteration 1, few-shot prompting was explored in addition to zero-shot. Temperature values explored for Llama and Qwen models were *{*0, 0.2, 0.4, 0.6, 0.8, 1.0*}* and *{*0, 0.4, 0.8, 1.2, 1.6, 2.0*}* for Gemini. The full set of explored LLM-derivation methods is outlined in Table 7.

### 9.2 Model Configuration Selection

The primary goal of the work described in this section was to identify a specific best configuration to use for each residual prediction setup being considered, separately for each response sample. All this work was carried out within the development set, and the resulting configurations were then pre-registered for deployment in the held-out set.

Final model configurations differ across response-samples (adolescent/parent) and residual setups. The method of deriving feature vectors from text is considered a pseudo-hyperparameter, and was chosen in the development set alongside *λ* ridge regression penalties. Note that *S*-layers utilize simple linear regression, whereas all *Q* - and *T*-layers employ penalized ridge regression. As such, the selection of LLM-derivation method was only relevant to setups involving *T*, and ridge penalties only relevant to setups involving *Q* or *T*.

Also note that pseudo-hyperparameters and ridge parameters were selected independently for each response-sample/residual-setup pair, since each combination presents a unique model. Let *λ_XY_* represent the regularization parameter assigned to layer *Y* in an *XY* setup. In an *SQT* model, distinct values of *λ* may have been selected for the *Q-*(*λ_SQ_*) and *T-*(*λ_SQT_*) layers. Additionally, the value of *λ_SQ_*may differ from that of *λ_Q_*, and *λ_SQT_* may differ from *λ_QT_*. Further, the LLM-derivation configuration may differ across *T*, *QT*, *ST*, and *SQT*, both within and across response-samples. The full search space is outlined in Table 7.

**Table 7:**
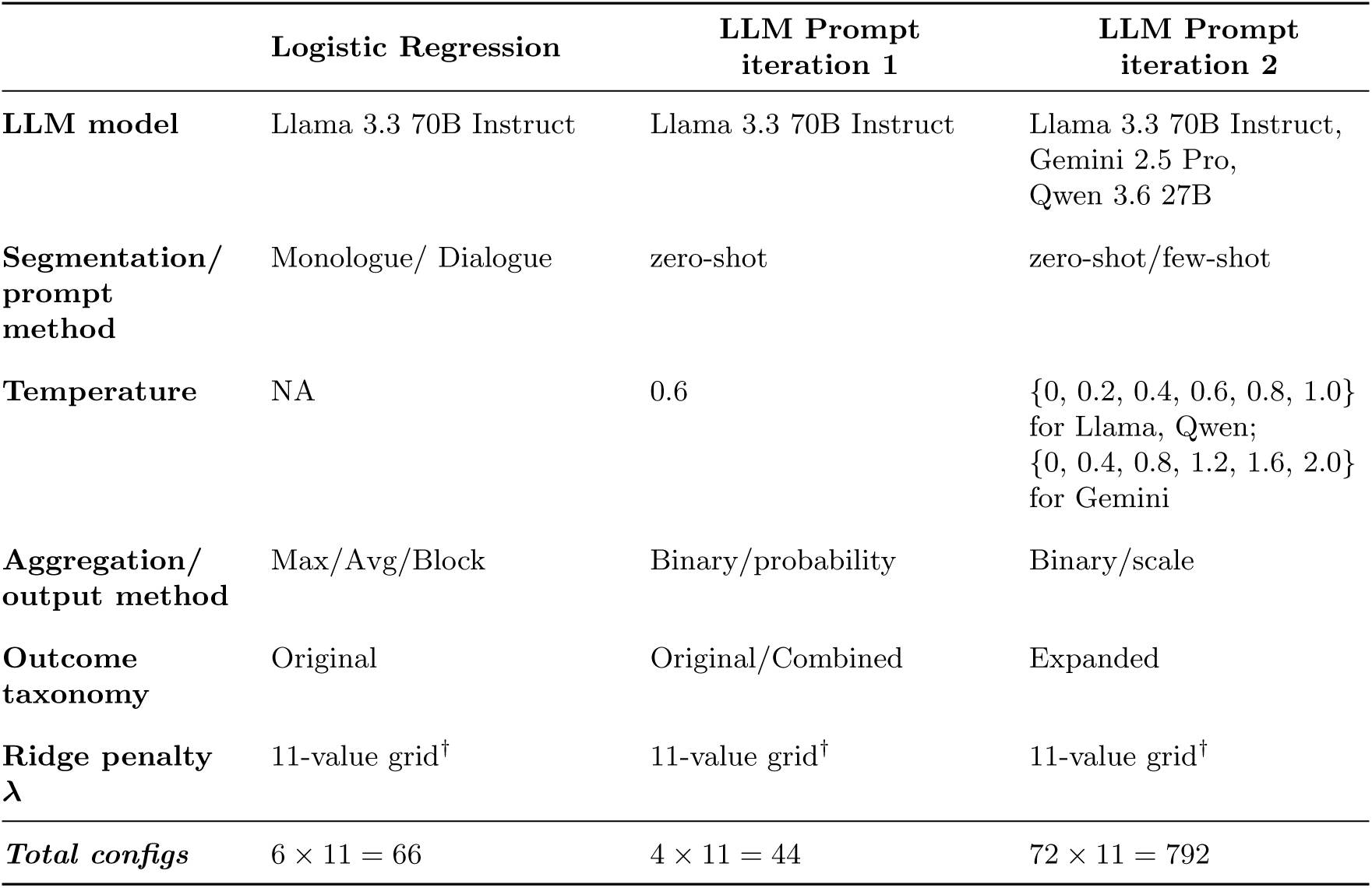
Text-derivation configurations explored at the *T* level, by family of settings. *^†^*{0.1, 0.5, 0.75, 1, 2.5, 5, 15, 25, 50, 75, 100}

#### 9.2.1 Performance Exploration

The performance of all possible configurations relevant to each residual setup was explored in the development set via 10-time repeated 3-fold cross-validation (CV). Each repeat utilized a different random seed, and the 3 folds were split stratified by *CAT-D* eligibility group. The primary metric of comparison was *repeat-aggregated R*^2^, which calculated *R*^2^ across concatenated out-of-fold (OOF) predictions within each repeat, then averaged across the 10 scores.

Note that all pseudo-hyperparameters (LLM-derivation method) of *T*-layers in *ST, QT*, and *SQT*, in addition to values for *T*-layer *λ*, were explored only in supplement to the best performing *Q* and *SQ* models. Thus, *λ_Q_*/*λ_SQ_* were frozen prior to exploration of all pseudo-hyperparameters and *λ_QT_* /*λ_SQT_* values. This ensured that signal from *T* supplements the best available item-level symptom-scale (*Q*) configuration, rather than compensating for a sub-optimally regularized *Q*-layer. Additionally, this allows for a consistent comparison of full/reduced model pairs like *SQT/SQ*. The *repeat-aggregated R*^2^ of each setup in parent- and adolescent-report samples are displayed in Table 8, along with their corresponding selected regularization and pseudo-hyperparameter values.

#### 9.2.2 Selection Framework/Criteria

There are no hyperparameters to select for *S*-layers. For *Q*, only *λ* regularization values were selected, chosen as those that resulted in the highest *repeat-aggregated R*^2^ performance in the development set. *T* - layers required the selection of both pseudo-hyperparameters and *λ*. For each selection of *T, ST, QT*, and *SQT*, configurations were ranked on *repeat-aggregated R*^2^ *scores*, the top 25 of which were compared for final selection.

Of the top models, further comparisons were made via *repeat-fold-aggregated R*^2^ score, which averages 30 OOF *R*^2^ results across each of the 3 folds in each of the 10 repeats. Although prone to more variance, this metric allows for comparison at a more granular scale. A second filtering step was performed among top models using the *baycomp* (Benavoli, Corani, Dem̌sar, & Zaffalon, 2017) package’s *two on single()* function, which performs a pairwise Bayesian correlated t-test, on the 30 paired *repeat-fold-aggregated R*^2^ observations across 2 given models, with the area of practical equivalence set to 0.01 (1% variance explained). The configuration with the highest *repeat-aggregated R*^2^ *score* was compared to each of the subsequent 24 within this framework. Configurations were retained for final consideration if the comparison was either conclusive in favor of equivalence (*P* (equal) *>* 0.5) or inconclusive (all of *P* (best better), *P* (equal), and *P* (best worse) falling below 0.5); that is, configurations were excluded only when the comparison conclusively favored one model over the other. Final models, displayed in Table 8, were selected from the surviving models with the following criteria, in order of importance:

1. No outlying residual error profile

- Configurations whose residuals did not cluster closely with those of the best-performing model were excluded. This was based on hierarchical clustering of pairwise residual correlations and their corresponding heatmaps.
2. Prompted LLM rather than LR

- Prompting allows for more reliable equivalence between what is intended to be assessed, and what is actually assessed
- No remaining choices across any configuration involved LR after this step
3. LLM model preferred in following order: Qwen, Llama, Gemini

- Qwen: open-weight, smaller than Llama
- Llama: open-weight
- Gemini: closed source
4. Prompt Method: few-shot, zero-shot

- Few-shot allows for a more reliable assessment of what is intended to be assessed. More is left to model biases in zero-shot
- Note that all remaining choices across all configurations only involved scale output type after this step. As a result, *output type* (binary, probability, scale) was not considered for selection criteria
5. Lambda Regularization: higher

- Higher regularization increases reliability
6. Temperature: lower

- More determinism increases reliability

**Table 8:** Final frozen model configurations and corresponding performance estimates, by model set-up and informant. *Repeat-aggregated R*^2^ is the development set cross-validation performance on which selection was based. Configurations selected during exploratory analysis in the develop-ment set only. All reported model configurations are unchanged from those presented in Table 4.

| **Set-up** | **Informant** | **LLM<br>Model** | **LLM<br>Prompt** | ***λ<sub>Q</sub>*** | ***λ<sub>T</sub>*** | **Temp.** | **Repeat-agg.<br>*R*<sup>2</sup>** |
|---|---|---|---|---|---|---|---|
| *S* | Parent | – | – | – | – | – | 0.182 |
|  | Adolescent | – | – | – | – | – | 0.123 |
| *Q* | Parent | – | – | 5 | – | – | 0.211 |
|  | Adolescent | – | – | 15 | – | – | 0.106 |
| *T* | Parent | Llama | few | – | 5 | 1.0 | 0.208 |
|  | Adolescent | Gemini | few | – | 5 | 0.4 | 0.145 |
| *SQ* | Parent | – | – | 15 | – | – | 0.226 |
|  | Adolescent | – | – | 50 | – | – | 0.141 |
| *ST* | Parent | Llama | few | – | 15 | 0.0 | 0.248 |
|  | Adolescent | Gemini | few | – | 25 | 0.4 | 0.191 |
| *QT* | Parent | Qwen | zero | 5 | 50 | 0.0 | 0.256 |
|  | Adolescent | Gemini | few | 15 | 5 | 0.4 | 0.207 |
| *SQT* | Parent | Qwen | zero | 15 | 75 | 0.0 | 0.264 |
|  | Adolescent | Gemini | few | 50 | 15 | 0.4 | 0.211 |

### 9.3 Step-down MinP Procedure

Within this procedure, p-value *p_j_* for sub-hypothesis *j* is calculated as:

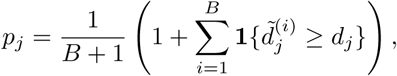

where *B* = 1000 is the number of permutation iterations, 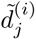 is the observed null difference at permutation iteration *i* for sub-hypothesis *j*, and *d_j_* is the true observed difference for sub-hypothesis *j*. In addition to observed *d_j_*, each 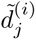 is also transformed into a p-value:

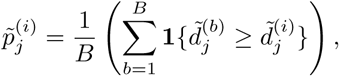

forming three distinct distributions of null p-values, one pertaining to each sub-hypothesis of 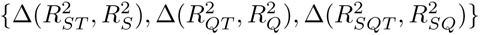. Note that the addition of 1 to the numerator and denominator is not included here because 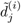 is included in the *B* null values being counted for hypothesis *j*.

The observed p-values are ordered from smallest to largest, *p*_(1)_ *≤ p*_(2)_ *≤ p*_(3)_, where *p*_(*k*)_ denotes the *k*-th smallest observed p-value in {*p*_1_*, p*_2_*, p*_3_}, and 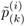 is the null p-value from iteration *i* for the sub-hypothesis with the *k*-th smallest observed p-value. Adjusted p-values are computed step-down. The null statistic for step *k* = 1 in each permutation iteration *i* is 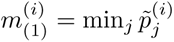: the minimum value across all three null p-values within permutation iteration *i*. The corresponding hypothesis of *p*_(1)_ is then removed from consideration for further testing. Accordingly, for step *k* = 2 in permutation iteration *i*, 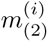 is the minimum value across the null p-distributions at iteration *i* of the two remaining sub-hypotheses 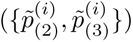. The remaining adjusted null distribution, *m*_(3)_, is simply the single remaining distribution of null p-values: *p̃*_(3)_. Using these step-specific null distributions, the step-wise p-value for the *k*-th ordered hypothesis is calculated as:

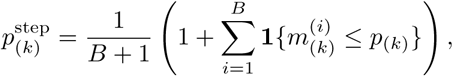

after which monotonicity is enforced by taking the cumulative maximum over preceding steps:

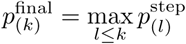

Because all three null differences within an iteration are computed from a single shared shuffle, the permutation distribution preserves the dependence among the hypotheses, so the procedure adapts to their correlation rather than assuming independence, serving to increase power over conservative alternative FWER controls such as the Bonferroni correction. Significance is set at the standard *α* = 0.05.

### 9.4 Supplemental Tables/Figures

**Table 9:**
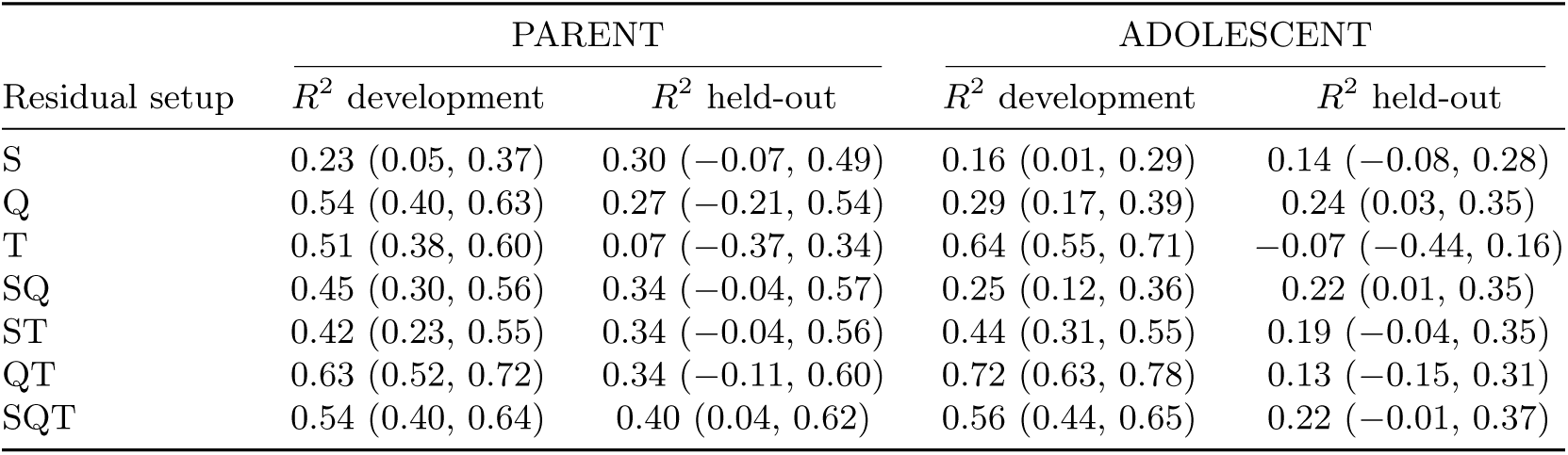
R^2^ (95% CIs) performance of models by residual setup and response-sample. Models fit on the development set only. Values are rounded to the nearest hundredth. *R*^2^ held-out values are unchanged from those in Table 5, and are presented here for reference.

**Table 10:**
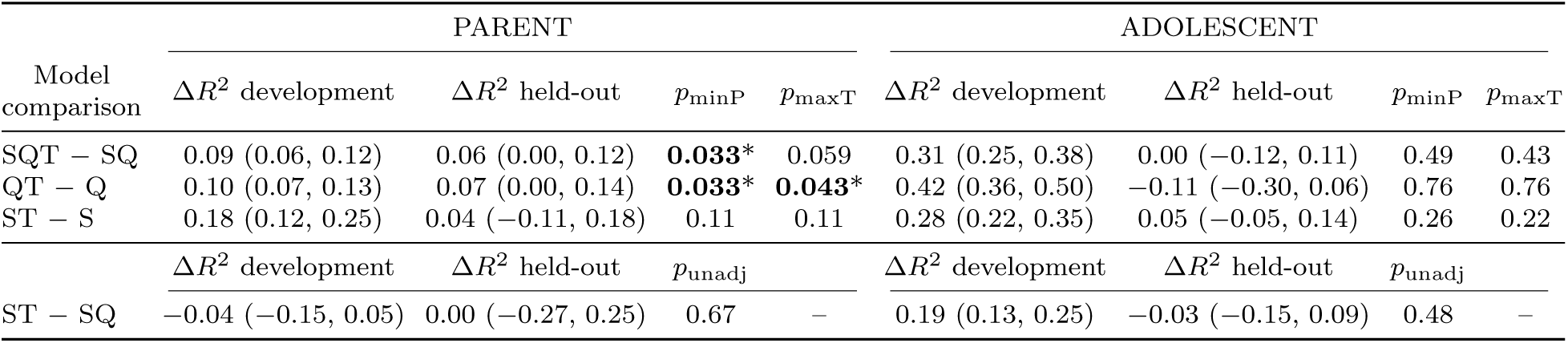
Differences (95% CIs) in *R*^2^ performance of models predicting depression impact in development and held-out sets in parent- and adolescent-report samples. Held-out set Δ*R*^2^ and all reported p-values are unchanged from those in Table 6, and are presented here for reference. Δ*R*^2^ values are rounded to the nearest hundredth. Models fit on the development set only. FWER-adjusted p-values from the Westfall– Young step-down minP (*p*_minP_) and maxT (*p*_maxT_) procedures are reported for *SQT/SQ, ST/S,* and *QT/Q*. Raw unadjusted p-values are reported for *ST/SQ*. Bold and * denote *p <* 0.05.

| Model comparison | PARENT |  |  |  | ADOLESCENT |  |  |  |
|---|---|---|---|---|---|---|---|---|
|  | Δ*R*<sup>2</sup> development | Δ*R*<sup>2</sup> held-out | *p*<sub>minP</sub> | *p*<sub>maxT</sub> | Δ*R*<sup>2</sup> development | Δ*R*<sup>2</sup> held-out | *p*<sub>minP</sub> | *p*<sub>maxT</sub> |
| SQT − SQ | 0.09 (0.06, 0.12) | 0.06 (0.00, 0.12) | **0.033**\* | 0.059 | 0.31 (0.25, 0.38) | 0.00 (−0.12, 0.11) | 0.49 | 0.43 |
| QT − Q | 0.10 (0.07, 0.13) | 0.07 (0.00, 0.14) | **0.033**\* | **0.043**\* | 0.42 (0.36, 0.50) | −0.11 (−0.30, 0.06) | 0.76 | 0.76 |
| ST − S | 0.18 (0.12, 0.25) | 0.04 (−0.11, 0.18) | 0.11 | 0.11 | 0.28 (0.22, 0.35) | 0.05 (−0.05, 0.14) | 0.26 | 0.22 |
|  | Δ*R*<sup>2</sup> development | Δ*R*<sup>2</sup> held-out | *p*<sub>unadj</sub> |  | Δ*R*<sup>2</sup> development | Δ*R*<sup>2</sup> held-out | *p*<sub>unadj</sub> |  |
| ST − SQ | −0.04 (−0.15, 0.05) | 0.00 (−0.27, 0.25) | 0.67 | – | 0.19 (0.13, 0.25) | −0.03 (−0.15, 0.09) | 0.48 | – |

**Table 11:**
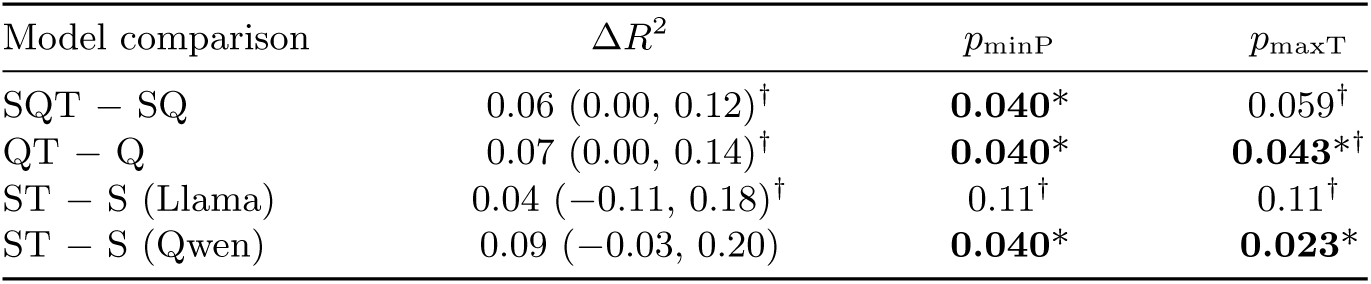
Differences (95% CIs) in held-out *R*^2^ for model comparisons in the parent-report sample. Comparisons include a post-hoc *ST* model built on Qwen (*Scale, Zero-shot, Temp=0.0, λ_T_*= 50), in addition to the original *ST* model built on Llama. Observed Δ*R*^2^ values for *SQT-SQ, QT-Q*, and Llama *ST-S* are unaffected by the addition of the Qwen *ST* model, but are included for reference. Additionally, their corrected p-values are subject to change with the additional Qwen *ST* hypothesis, as FWER-adjusted p-values from the Westfall–Young step-down minP (*p*_minP_) and maxT (*p*_maxT_) procedures are employed with all four hypotheses. Bold and * denote *p <* 0.05. *†* denotes values are unchanged from confirmatory analysis.

| Model comparison | Δ*R*<sup>2</sup> | *p*<sub>minP</sub> | *p*<sub>maxT</sub> |
|---|---|---|---|
| SQT − SQ | 0.06 (0.00, 0.12)<sup>†</sup> | **0.040**\* | 0.059<sup>†</sup> |
| QT − Q | 0.07 (0.00, 0.14)<sup>†</sup> | **0.040**\* | **0.043**\*<sup>†</sup> |
| ST − S (Llama) | 0.04 (−0.11, 0.18)<sup>†</sup> | 0.11<sup>†</sup> | 0.11<sup>†</sup> |
| ST − S (Qwen) | 0.09 (−0.03, 0.20) | **0.040**\* | **0.023**\* |

**Table 12:** DAWBA depression impact items used to construct the aggregated impact score for adolescent self-responses. Parent items are similar, with instances of *‘you’* and *‘your’* replaced with *‘him or her’* and *‘his or her’*, in reference to the parent’s adolescent. Response options were: 0 = Not at all, 1 = A little, 2 = A medium amount, 3 = A great deal.

| Item | Content | Range |
|---|---|---|
| H19 | How much has your sadness, irritability or loss of interest upset or distressed you? | 0–3 |
| *H20 Has your sadness, irritability or loss of interest interfered with…* |  |  |
| H20a | how well you get on with the rest of the family? | 0–3 |
| H20b | making and keeping friends? | 0–3 |
| H20c | learning or class work? | 0–3 |
| H20d | playing, hobbies, sports or other leisure activities? | 0–3 |
| Aggregated impact score |  | 0–15 |

**Figure 3:**
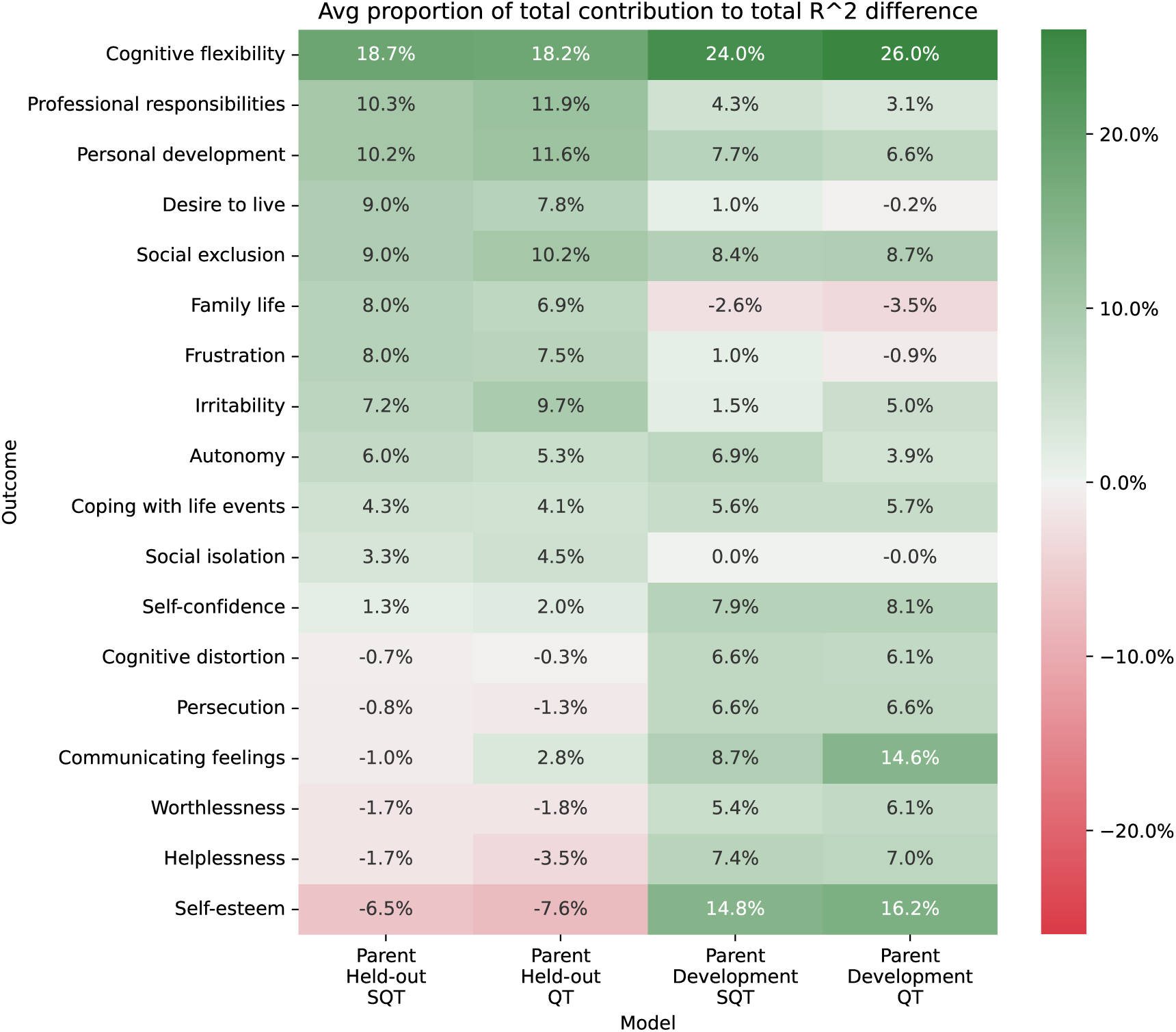
Aggregated normalized increment in % variance explained (*R*^2^) when an outcome is added to the model (one of *QT* or *SQT* in the parent-response sample). Displayed values are averaged over out-of-sample performance increments across 2000 orderings in the held-out set, and mean out-of-fold performance increments across 2000 orderings and 10-time repeated 3-fold CV (30 folds) in the development set, shown as the proportion of total incremental contribution of *T* in their respective setup.

#### 9.4.1 Free-text Questions

The following is the set of 10 free-text questions asked to adolescents in the DAWBA (Goodman et al., 2000).

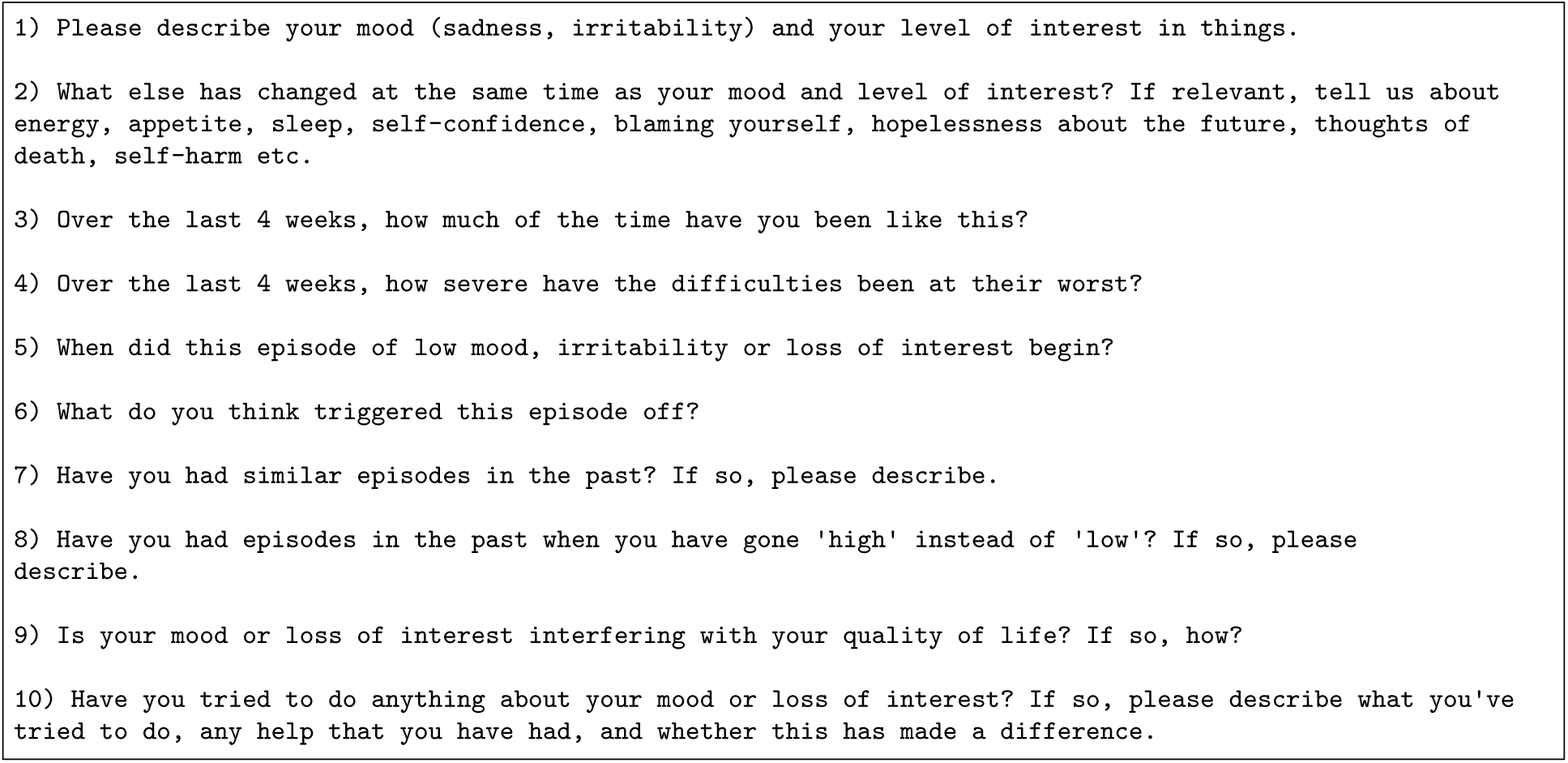

Note that the same questions are given to parents, with instances of ‘*you*’ or ‘*your*’ replaced with ‘*he or she*’ or ‘*his or her*’, in reference to the parent’s adolescent. For example, question 1 for the parent-response sample is: *Please describe his/her mood (sadness, irritability) and his or her level of interest in things*.

#### 9.4.2 Example Prompt

The following is an example zero-shot scale prompt for the *irritability* outcome of a parent-reported free-text response. Only two question/answer pairs are included here, but actual prompts contain all 10, even if some (up to 5) are left “Not answered”.

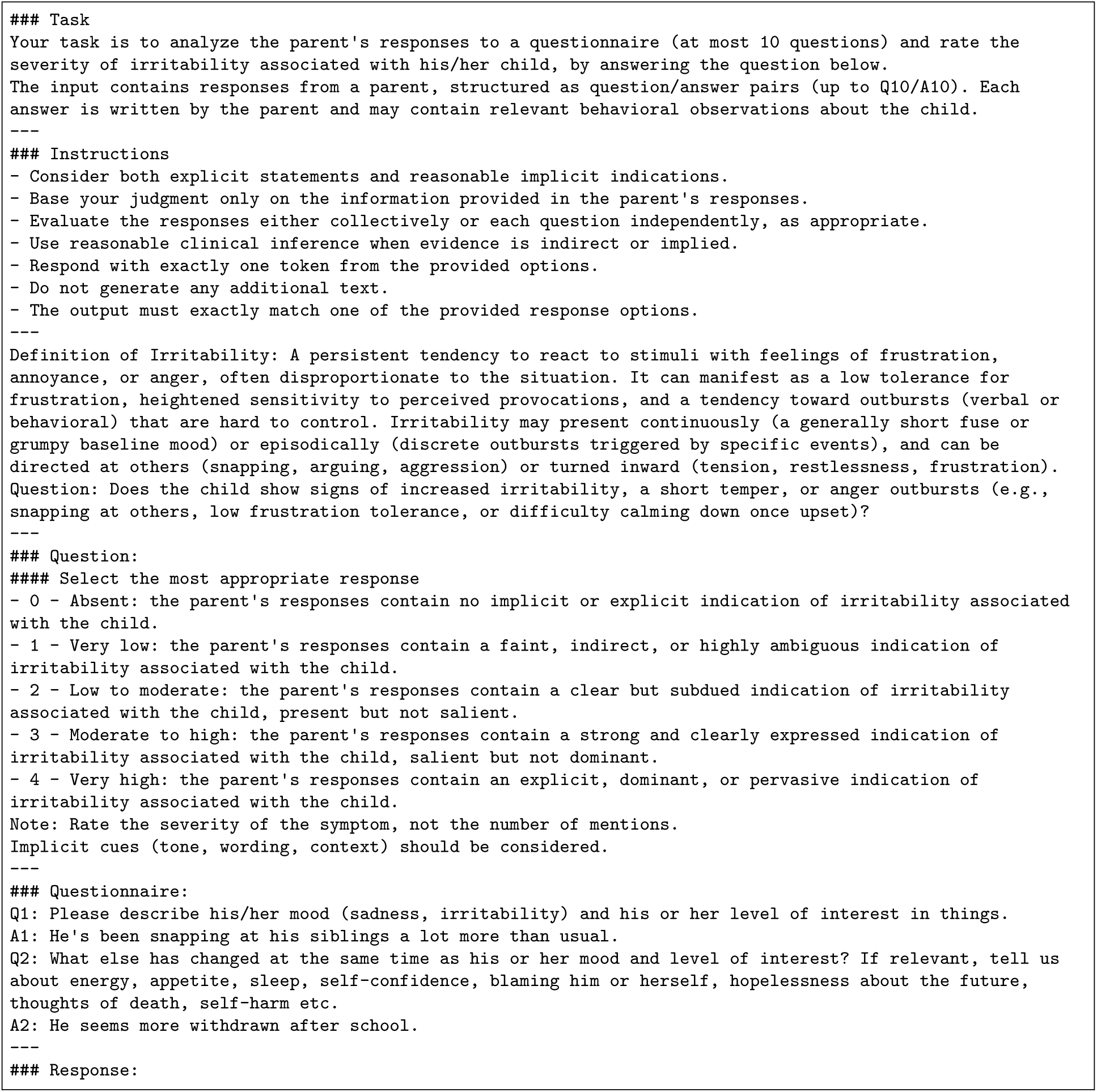

|  |
|---|
| ### Task<br>Your task is to analyze the parent's responses to a questionnaire (at most 10 questions) and rate the severity of irritability associated with his/her child, by answering the question below.<br>The input contains responses from a parent, structured as question/answer pairs (up to Q10/A10). Each answer is written by the parent and may contain relevant behavioral observations about the child.<br>---<br>### Instructions<br>- Consider both explicit statements and reasonable implicit indications.<br>- Base your judgment only on the information provided in the parent's responses.<br>- Evaluate the responses either collectively or each question independently, as appropriate.<br>- Use reasonable clinical inference when evidence is indirect or implied.<br>- Respond with exactly one token from the provided options.<br>- Do not generate any additional text.<br>- The output must exactly match one of the provided response options.<br>---<br>Definition of Irritability: A persistent tendency to react to stimuli with feelings of frustration, annoyance, or anger, often disproportionate to the situation. It can manifest as a low tolerance for frustration, heightened sensitivity to perceived provocations, and a tendency toward outbursts (verbal or behavioral) that are hard to control. Irritability may present continuously (a generally short fuse or grumpy baseline mood) or episodically (discrete outbursts triggered by specific events), and can be directed at others (snapping, arguing, aggression) or turned inward (tension, restlessness, frustration).<br>Question: Does the child show signs of increased irritability, a short temper, or anger outbursts (e.g., snapping at others, low frustration tolerance, or difficulty calming down once upset)?<br>---<br>### Question:<br>#### Select the most appropriate response<br>- 0 - Absent: the parent's responses contain no implicit or explicit indication of irritability associated with the child.<br>- 1 - Very low: the parent's responses contain a faint, indirect, or highly ambiguous indication of irritability associated with the child.<br>- 2 - Low to moderate: the parent's responses contain a clear but subdued indication of irritability associated with the child, present but not salient.<br>- 3 - Moderate to high: the parent's responses contain a strong and clearly expressed indication of irritability associated with the child, salient but not dominant.<br>- 4 - Very high: the parent's responses contain an explicit, dominant, or pervasive indication of irritability associated with the child.<br>Note: Rate the severity of the symptom, not the number of mentions.<br>Implicit cues (tone, wording, context) should be considered.<br>---<br>### Questionnaire:<br>Q1: Please describe his/her mood (sadness, irritability) and his or her level of interest in things.<br>A1: He's been snapping at his siblings a lot more than usual.<br>Q2: What else has changed at the same time as his or her mood and level of interest? If relevant, tell us about energy, appetite, sleep, self-confidence, blaming him or herself, hopelessness about the future, thoughts of death, self-harm etc.<br>A2: He seems more withdrawn after school.<br>---<br>### Response: |

